# Effect of 2021 Assembly Election in India on Covid-19 Transmission

**DOI:** 10.1101/2021.05.30.21258040

**Authors:** Souvik Manik, Sabyasachi Pal, Manoj Mandal, Mangal Hazra

## Abstract

India is one of the countries in the world which is badly affected by the Covid-19 second wave. Assembly election in four states and a union territory of India was taken place during March-May 2021 when the Covid-19 second wave was close to its peak and affected a huge number of people. We studied the impact of assembly election on the effective contact rate and the effective reproduction number of Covid-19 using different epidemiological models like SIR, SIRD, and SEIR. We also modeled the effective reproduction number for all election-bound states using different mathematical functions. We separately studied the case of all election-bound states and found all the states shown a distinct increase in the effective contact rate and the effective reproduction number during the election-bound time and just after that compared to pre-election time. States, where elections were conducted in single-phase, showed less increase in the effective contact rate and the reproduction number. The election commission imposed extra measures from the first week of April 2021 to restrict big campaign rallies, meetings, and different political activities. The effective contact rate and the reproduction number showed a trend to decrease for few states due to the imposition of the restrictions. We also compared the effective contact rate, and the effective reproduction number of all election-bound states and the rest of India and found all the parameters related to the spread of virus for election-bound states are distinctly high compared to the rest of India.

## 1 Introduction

In November 2019, extremely infectious Corona Virus Disease 2019 (COVID-19) was found in the Wuhan province of China^1^. Wuhan Municipal Health Commission notified the confirmation of identification of COVID-19 on December 31, 2019^2^. The name of the Coronavirus disease announced officially as COVID-19 by World Health Organisation (WHO) and the International Committee on Taxonomy of Viruses (ICTV) also announced the name of the virus as Severe Acute Respiratory Syndrome Corona Virus 2 (SARS-CoV-2) on February 11, 2020^3^. Presently, four mutant variants of Coronavirus are dominant i.e. a) UK Variant [VOC 202012/01 (B.1.1.7)] b) South Africa variant [501Y.V2 (B.1.351)] c) Brazil variant [P.1 (P.1)] [11] and d) Indian variant [B.1.617] [9]. WHO warned that the Indian variant, B.1.617, is spreading rapidly in India, making up more than 28% of samples from positive tests. Indian strain (B.1.617) appears more contagious than other variants of SARS-CoV2^4^.

India enrolled the first new positive case of Novel Coronavirus patient of Kerala in January 2020 [21]. Till May 2, 2021, 20 million total cases were reported with 0.37 million daily new cases and 0.21 million total death^5^. To control the spread of the virus, few lockdowns were strictly imposed throughout India from March 24, 2020, and later, India started to reopen in phases from June 2020 [12]. COVID vaccination for people of age 50 years or above started in India on January 16, 2021^6^ which was later opened up for the lower age group^7^.

The second wave of COVID-19 began around February 11, 2021, in India. The effect of the second wave was severe with a record high daily cases of nearly three times compared to the peak of the first wave [17]. The daily deceased cases near the peak of the second wave were also nearly three times than the first wave peak, which has a great impact on society. The double mutant B.1.1.7 variant was mainly responsible for the second wave in India. In India, assembly elections took place for four states and a union territory in multiple phases during the pandemic situation (March-May 2021) when the Covid-19 second wave was close to its peak and affected a huge number of people. On February 26, 2021, the Election Commission of India (ECI) announced the tentative date of polling in different states. Polling in Assam and West Bengal was started on March 27, 2021, in three phases and eight phases respectively. Polling in Kerala and Tamil Nadu was in a single phase on April 6, 2021. The election result was announced on May 2, 2021^8^,^9^. Election procedure with 186.8 million electors were taken place using 0.27 million polling stations in Tamil Nadu, West Bengal, Kerala, and Assam. Big campaigns and many huge rallies took place in the election-bound regions, often with hundreds of thousands of people without following strict guidelines to restrict the spread of the Coronavirus which result in a rapid increase in infected individuals and casualties [1]. It is well established that overcrowded gatherings increased the transmission rate of the virus drastically. Noting the impact of the election on the spread of the virus, the Madras High Court reproofed the ECI for approval of political rallies^10^. After the reprieval by Madras High Court, on April 9, 2021, ECI imposed few restrictions during the time of campaigns, election and counting to control spread of the Coronavirus [18]. Noting the surge of the virus, ECI announced more restrictions on political activities and the counting process of the election on April 27 and 28, 2021 [10, 3].

In this paper, we studied the variation of various driving parameters of the pandemic using different mathematical models and try to understand the effect of the pre-election activities and the impact of multiple phases of election in few states. We investigated the variation of effective contact rate (*β*_*n*_) and effective reproduction number (ℝ (t)) of Coronavirus disease using different mathematical models for election and non-election bound states during the assembly election of India in 2021.

We have summarised basic mathematical models for epidemics used in the article in Section 2. The data analysis and methodology are discussed in Section 3. The results of our study are given in Section 4. The discussion and conclusion are summarised in the Section 5 and 6 respectively.

## 2 Mathematical Models

We have used different mathematical models to study the effect of the election on the spread of Coronavirus. The key element of these models is to calculate the basic reproduction number (ℝ_0_). The average number of people affected by a single infected person over time is represented by ℝ_0_. ℝ_0_ *>* 1 implies that the spread of the disease is increasing, ℝ_0_ = 1, represents that the spread is stable and ℝ_0_ *<* 1, means the spread is expected to stop over a time. Using different models, we investigated the evolution of the effective reproduction number ℝ(*t*) for various election-bound states [15].

### 2.1 SIR Model

There are several models for studying contagious disease transmission in a large population. The simplest compartmental model that can explain the evolution of an outbreak at the population level is the Susceptible-Infected-Recovered (SIR) model [22, 2, 15]. This model was used to study various time-dependent parameters such as recovery rate (*γ*), contact rate (*β*), and effective reproduction number (ℝ). At any given time *t, S*(*t*) (total population — confirmed), *I*(*t*) (confirmed — recovered — deaths), and ℝ(*t*)) (recovered + deaths) be the total number of susceptible individuals, infected individuals, and recovered individuals respectively. *N* is the total population, which is assumed to be constant during our study.

These parameters can be expressed as in the form of fractions: 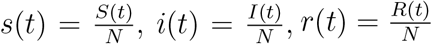

where, *s*(*t*)+*i*(*t*)+*r*(*t*)=1 from the conservation law

SIR model can be expressed by following set of differential equations.

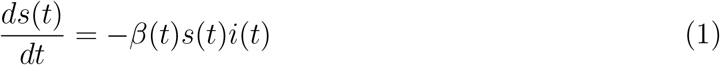

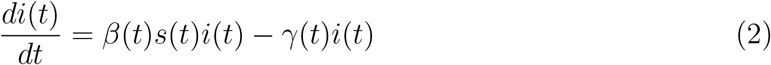

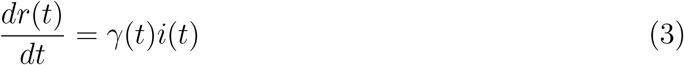

We can write from equation 2,

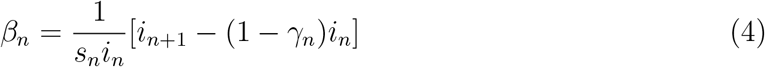

The effective reproduction number ℝ(t) can be figured out from this equation.

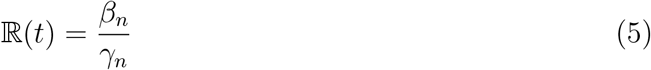

We studied the variation of recovery rate (*γ*_*n*_), contact rate (*β*_*n*_), and effective reproduction number (ℝ(*t*)) of Coronavirus which are dependent on time.

### 2.2 SIRD Model

The Susceptible-Infectious-Recovered-deceased (SIRD) [2, 15] is modified version of the SIR model (Section 2.1). In this model, there is a separate compartment for the deceased and recovered individuals. The time-dependent function *S*(*t*), *I*(*t*), *R*(*t*) and *D*(*t*) be the total number of susceptible individuals, the total number of infected individuals, the total number of recovered individuals and the total number of deceased individuals respectively and *N* is the total population, which is assumed to remain constant over the study. Using this model we studied time dependence functions like the recovery rate (*γ*_*n*_), contact rate (*β*_*n*_), mortality rate (*α*_*n*_) and effective reproduction number (ℝ(*t*)).

*s*(*t*), *i*(*t*), *r*(*t*) and *d*(*t*) can be expressed in fractional form: 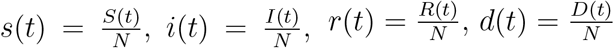

where, *s*(*t*) + *i*(*t*) + *r*(*t*) + *d*(*t*) = 1

SIRD model can be expressed by following set of differential equations.

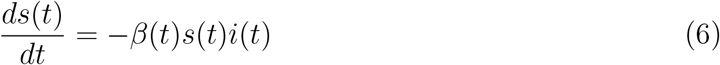

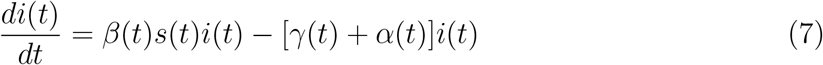

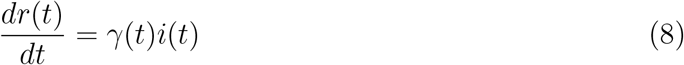

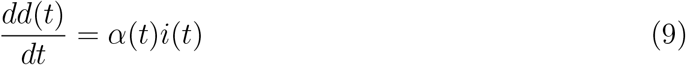

From the equation 7 we may write

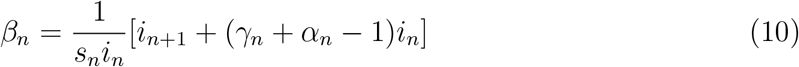

The effective reproduction number ℝ(t) can be expressed as

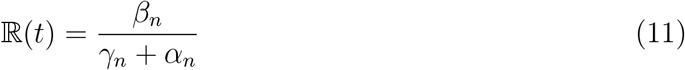

### 2.3 SEIR Model

The most studied epidemic model is the Susceptible-Exposed-Infectious-Removed (SEIR) model [16, 15, 5, 7]. The SEIR model is a variation of the SIR model (Section 2.1). We used the average incubation period 1*/σ* (=7) as a constant in the SEIR model. The time-dependent function *S*(*t*), *E*(*t*), *I*(*t*) and *R*(*t*) be the total number of susceptible individuals, the total number of exposed individuals, the total number of infected individuals, and the total number of removed individuals (recovered or deceased) respectively. We assumed the total population (*N*) remains constant during the study. Here, *γ*_*n*_ is the recovery rate, *β*_*n*_ and ℝ(*t*) are the effective contact rate, and effective reproduction number of Coronavirus respectively. The contact rate (*β*) is defined as the product of the average number of contacts per person per unit time by the probability of disease transmission in contact between susceptible and infectious individuals and *γ* is the recovery rate. The introduction of the exposed individuals (*E*) in the SEIR model makes the model more efficient than the SIR model.

*s*(*t*), *e*(*t*), *i*(*t*) and *r*(*t*) can be expressed in fractional form:

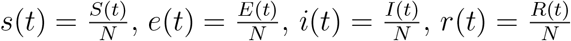

From the conservation law, we may write: *s*(*t*) + *e*(*t*) + *i*(*t*) + *r*(*t*) = 1

The SEIR model can be expressed by following set of differential equations

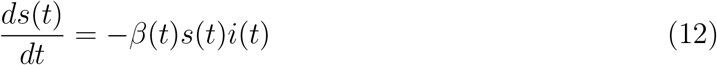

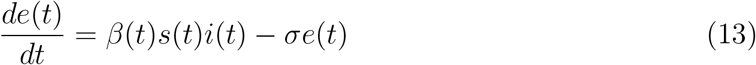

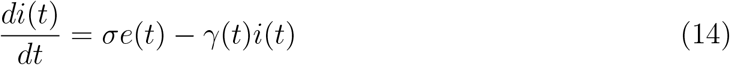

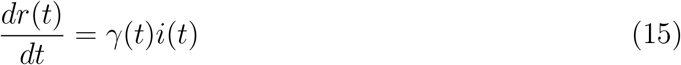

By combining eqn 13 and 14,

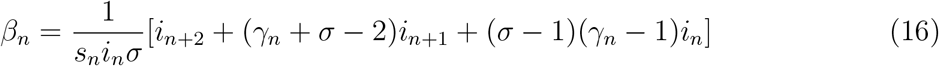

The effective reproduction number ℝ(t) can be expressed from this equation.

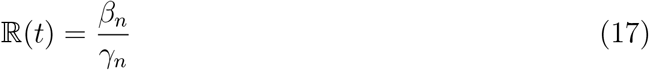

### 2.4 Time Dependent Exponential *β* Suppression Model

As the pandemic evolved, various control measures [8] were introduced, including lock-down, social distancing, and increased hygiene, causing transmission coefficients *β* to become time dependent [13]. Apart from this, the drop in the susceptible population also decreases *β* [14, 19, 4, 20].

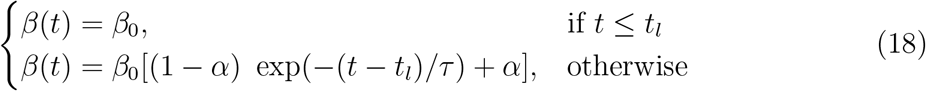

where

*β*_0_ = The initial contact rate

t_*l*_ = A certain time for starting lockdown

*µ* = 1/*τ* = Decreasing rate or decay rate

*α* =*β*_*min*_/*β*_*max*_

In this model, *β*(*t*) varies from some constant *β*_0_ to *β*_0_(1 − *α*) between time *t* = *t*_*l*_ to *t* = ∞. From equation (18),

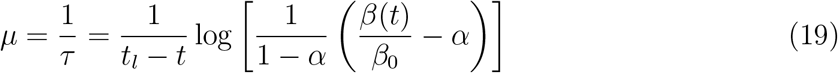

We used this model to fit the data of infected and recovered cases for election-bound states and summarized the fitting parameters in Table 2.

## 3 Data Analysis and Methodology

We used the COVID-19 data maintained by the Center for Systems Science and Engineering (CSSE) at Johns Hopkins University to track reported cases of COVID-19 in real-time [6]. The time-series data of India and state-wise can be accessed from these sources^11^,^12^. To compare the effect of the election on the spread of Coronavirus, we used time-series data of pre-election time and election time. We studied the impact of the election on the spread of the Coronavirus from February 2, 2021, to May 11, 2021.

We studied the variation of confirmed, recovered, and deceased individuals for all individual election-bound states and the cumulative effect of all election-bound states and rest of the India. The effective contact rate (*β*(*t*)), recovery rate (*γ*(*t*)), and effective reproduction number (ℝ(*t*)) were estimated from the mathematical equations discussed in Section 2 for whole India and all election-bound states separately. We also studied the evolution of these parameters using times series data.

To minimize the effect of noise, we used a three-day rolling mean variation for our study. We have calculated the values of transmission coefficients using three basic models (2.1, 2.2, 2.3) for four election-bound states (Assam, Kerala, Tamil Nadu, and West Bengal). To compare the effect of the election on the spread of Coronavirus, we studied separately cumulative statistics of all election-bound states and the rest of India.

We studied the variation of confirmed, recovered, and deceased individuals for all election-bound states, the cumulative effect of all election-bound states and the rest of India. The effective contact rate (*β*(*t*)), recovery rate (*γ*(*t*)), and effective reproduction number (ℝ(*t*)) were estimated from the mathematical equations discussed at Section 2 for whole India and all election-bound states separately. We also studied the evolution of these parameters using time series data.

The assembly election in the Union Territory of Puducherry was also held along with elections of four states. The election took place in a single phase on April 6, 2021^13^. The population of Puducherry is ∼1.7 million^14^, which is comparatively low than other election-bound states in India. So, we exclude Puducherry in our study.

## 4 Result

We studied the impact of the election, political activities, and a bulk amount of gatherings without maintaining Covid-19 protocols on the spread of Coronavirus. We used different basic mathematical models like SIR, SIRD, and SEIR to study the variation of different fundamental parameters related to a pandemic. Figure 1 showed the variation of different driving parameters of the pandemic over the whole of India. The first panel of Figure 1 showed the variation of confirmed, recovered, and deceased individuals, which indicated that the second wave of Coronavirus was effective from the second week of February 2021. The second panel of Figure 1 showed the variation of effective contact rate *β*(t) using SIR, SIRD, and SEIR models over the same time. *β*(t) showed a trend to increase from March 2021 and the value of *β*(t) varied between 0.05 and 0.3 and attained a maximum of ∼0.3 during the mid-week of March for the SEIR model. SIR and SIRD models showed a similar trend and varied between 0 and 0.17 and reached a maximum near March 19, 2021. The third panel showed the variation of recovery rate *γ*(t), which was varied between ∼0.06 to 0.12 during February–April. The variation of effective reproduction number showed in the bottom panel of Figure 1, which increased drastically during February–April for all the mathematical models. Using the SEIR model, the effective reproduction number (ℝ(*t*)) reached the highest value of ∼4 during the mid-week of March 2021. For SIR and SIRD models, the value of the effective reproduction number varied between 0.75 and 2.1 and peaked near March 28, 2021. For all three models, there was a sharp rise in ℝ(*t*) from March 27 which peaked near April 11, 2021. From Section 4.5, it will be evident that the increase in ℝ(*t*) is mostly contributed by four election-bound states.

**Figure 1:**
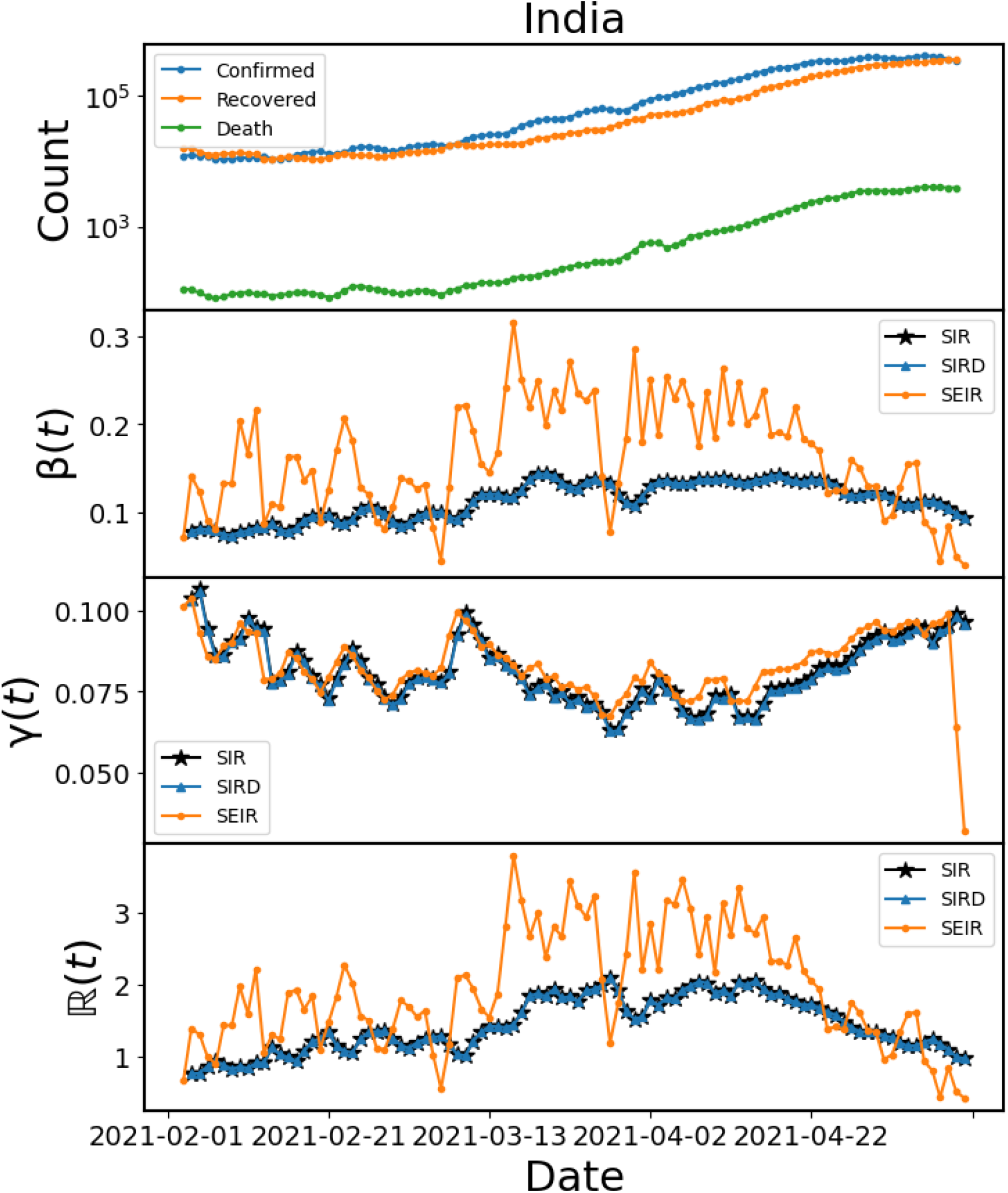
First panel: The variation of three types of Covid-19 affected populations (confirmed, recovered, and deceased individuals) with time. Second panel: Using Different mathematical models (SIR, SIRD, SEIR), the change in the number of contact rate (*β*(*t*)) with time. Third panel: Variation of recovery rate (*γ*(*t*)) with time using three models. Fourth panel: The variation of the effective reproduction number (ℝ(*t*)) for three models.

### 4.1 Kerala

The population of the Indian state of Kerala is ∼34.8 million in the year 2021^15^. The election in Kerala was held in only one phase. The variation of different parameters was shown in top left of Figure 2. The figure showed that due to single-phase elections, the spread of Coronavirus was significant but less compared to other states where the election was held in multiple phases. The number of infected, deceased individuals started to increase a week before the election when campaigns and rallies were at their peaks. The value of effective reproduction number was below 1.05 on the date of announcement of the election and increased to nearly 3.46, 1.45, and 1.46 for the SEIR, SIR, and SIRD models respectively after the election (on April 6) which continue to rise drastically even after the election. During the last week of April (on April 20), the reproduction number reached ∼13 for the SEIR model. The SIR and SIRD models also showed a rise in the value of effective reproduction number during the last week of April 2021 and reached the highest value of ∼5.

**Figure 2:**
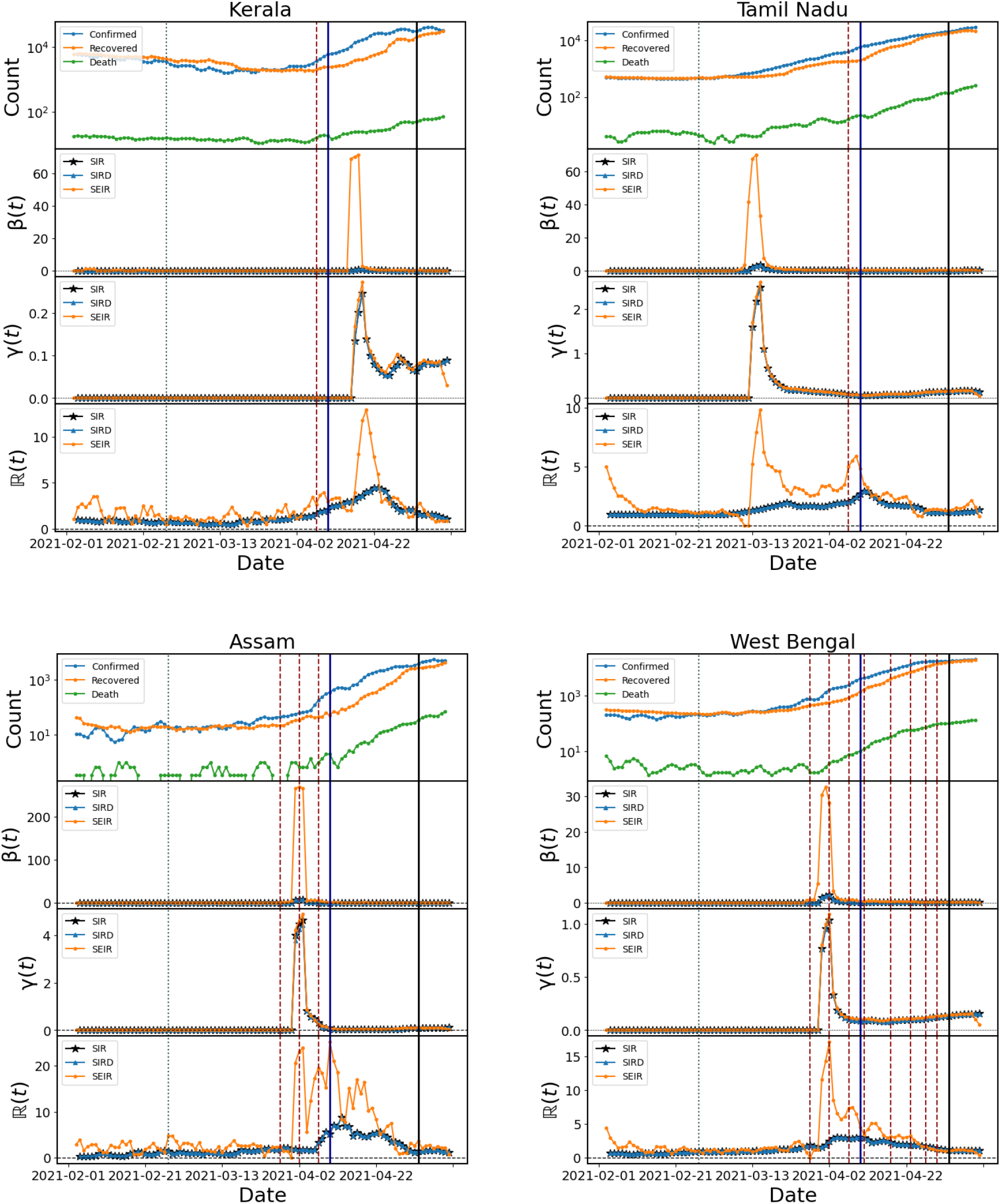
First panel: The standard variation of confirmed, recovered, and deceased individuals with time. Second panel: Variation of effective contact rate (*β*(*t*)) with time using SIR, SIRD and SEIR models. Third panel: Variation of recovery rate (*γ*(*t*)) with time for three basic models. Fourth panel: Variation of the effective reproduction number (ℝ(*t*)) using three models. The grey dotted lines indicated the announcement date of the election and red dashed lines pointed to the different phases of election for mentioned states. The blue solid line shows the date of warning notice of ECI to political parties. The black solid line indicates the date of counting and election result.

### 4.2 Tamil Nadu

The population of the Indian state of Tamil Nadu is ∼84.9 million in the year 2021^16^. The right top of Figure 2 showed the variation of different parameters related to the spread rate of Covid-19 for the state Tamil Nadu. The election of this state occurred in a single phase during the first week of April 2021.

The Figure implied that the effective reproduction number reached a peak during the second week of March 2021 when the election campaigns were in full swing. The effective reproduction number reached a value of 9.85 for the SEIR model during the third week of March 2021. There was another peak (∼5.9) in the effective reproduction number just after the date of the election. The value of the effective reproduction number was ∼1.08 on the date of announcement of the election (February 26, 2021) and increased drastically to a value of 5.9 for the SEIR model just after the election. The SIR and SIRD models also showed a trend of rising in the effective reproduction number just after the election and reached a maximum value of ∼3. The effective reproduction number started to decrease after the end of the election.

### 4.3 Assam

The population of the Indian state of Assam is ∼36.5 million in the year 2021^17^. The bottom left of Figure 2 showed the variation of different fundamental parameters of the pandemic for the state Assam. The red dotted vertical lines indicated the phases of the election of Assam. The number of daily infected individuals showed a sharp rise from the first phase of the election in Assam. The effective contact rate and the effective reproduction number with the SEIR model reached a peak after the second phase of the election (April 3, 2021). All the models showed a very high value of the effective reproduction number during all phases of the election which started to decrease after the last phase of the election was over and reached a low value after counting of the election was over. The value of effective reproduction number was 1.62 on the date of announcement of election which increased to 23.9 for the SEIR model after the second phase of election (on April 2, 2021) and increased drastically to a maximum value of ∼25 for SEIR model after the election (on April 10, 2021). The effective reproduction number showed a trend of rising from just after the election for SIR and SIRD models and reached a maximum value of ∼9.

### 4.4 West Bengal

For four election-bound states, we studied separately the impact of the election on the spread of Coronavirus. Indian state West Bengal is the only state where the election was held for a long duration of 33 days, with 8 phases which increased the spread rate of the virus drastically. In the bottom right of Figure 2, the variation of different parameters using 3 different mathematical models for West Bengal is shown. The population of West Bengal is ∼101.8 million in the year 2021^18^. The red dotted vertical lines show the time of different phases of the election. The date of announcement of the election and poll counting are also added to the Figure.

The Figure shows that the number of individuals infected and deceased increased sharply during the election phases and reached a peak near the second phase of the election when campaigns were at their peaks for all major political parties. Before the beginning of the election (on March 27, 2021), the daily confirmed cases were 812, which increased to 1733 after the second phase of the election (on April 2, 2021). One of the driving factors of a pandemic is the effective reproduction number, which was ∼0.95 on the date of announcement of the election and increased to a record value of nearly 17.14 for the SEIR model after the second phase of the election (on April 2, 2021). The effective reproduction number also showed a trend to increase for both the SIR and SIRD models from the first phase of the election. Before each phase of the election, a small but significant peak was visible in the reproduction number. The effective reproduction number decreased after all election-bound activity was over with the counting of the election.

### 4.5 Comparison of Spread Rate Between Election and Non-Election States in India

To compare the effect of the election on the spread of Coronavirus between election-bound states and the rest of India, we studied the cumulative statistics of all election-bound states and the rest of India. Figure 3 showed the variation of infected, recovered, and deceased individuals along with the effective contact rate, and the effective reproduction number for cumulative statistics of election-bound states (left) and the rest of India (right). The number of confirmed and deceased individuals was increased drastically during the election (as shown in the shaded region of Figure 3). The effective contact rate and effective reproduction number were showed a sharp rise for the cumulative study of all election-bound states during the time of election. The effective reproduction number was also increased drastically from early April 2021 and reached a maximum value of ∼8 for the SEIR model. For SIR and SIRD models, the peak value of ℝ(*t*) reached ∼2.5. The value of ℝ(*t*) was ∼1.03 on the date of announcement of the election and increased drastically to a record value of 8.3 for the SEIR model during election time (on April 11, 2021). The right side of Figure 3 showed the variation of different parameters for non-election states of India, which indicated that the value of the effective contact rate and effective reproduction number during the same time is less compared to election-bound states. So, it was evident that the impact of the election and political activities on the spread rate of the Coronavirus was significant for India.

**Figure 3:**
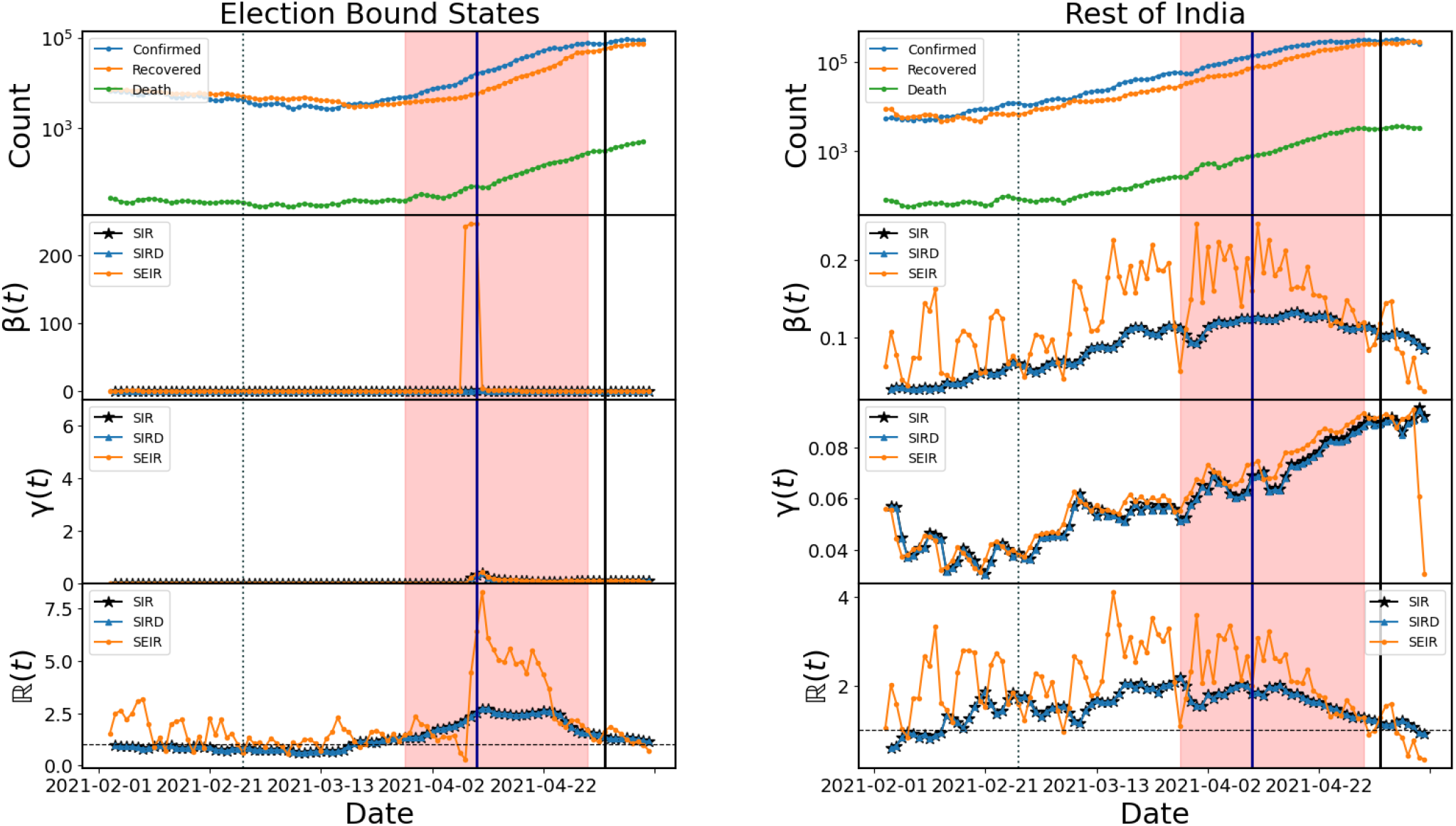
First panel: Variation of three days rolling mean of confirmed, recovered, and deceased individuals. Second panel: Variation of effective contact rate (*β*(*t*)) with time using SIR, SIRD, and SEIR models. Third panel: Variation of recovery rate (*γ*(*t*)) with time using these models. Fourth panel: Evolution of the effective reproduction number (ℝ(*t*)) using three models. The announcement date of the election is indicated by the grey dotted line and the announcement date of warning notice of ECI to political parties is shown by the blue solid line. For both plots, the pink shaded region referred to the time between starting and ending dates for the election in election-bound states. The black solid line shows the date of counting and election result.

### 4.6 Mathematical Modelling of Variation of Reproduction Number

We calculated the transmission coefficients and effective reproduction number using different mathematical models (SIR, SIRD, SEIR) for cumulative statistics of all election-bound states. During the election time (as shown in the shaded region of Figure 3), we fitted the effective reproduction numbers for all election-bound states with different mathematical functions i.e. Gaussian, Lorentzian, and Split-Lorentzian with the least square method for different epidemiological models (SIR, SIRD, and SEIR). Values of all parameters are summarised in Table 1.

**Table 1:** A summary of the fitting parameters for the models used to fit effective reproduction number during the election period.

| Model | Epidemiological Models | $A, \mu, \sigma, \sigma_r$ |
| --- | --- | --- |
| Gaussian | SIR | 94.84, 19.03, 14.50, — |
|  | SIRD | 94.84, 19.03, 14.50, — |
|  | SEIR | 108.54, 18.53, 7.54, — |
| Lorentzian | SIR | 148.85, 19.27, 17.91, — |
|  | SIRD | 148.85, 19.27, 17.91, — |
|  | SEIR | 150.37, 18.15, 7.90, — |
| Split Lorentzian | SIR | 143.72, 23.29, 23.61, 10.72 |
|  | SIRD | 143.72, 23.29, 23.61, 10.72 |

Figure 4, showed the variation of infected and removed individuals for all four election-bound states starting the date of the first election. The fitting with the SIR model with the time-dependent exponential *β* suppression model was shown for all four election-bound states (Kerala, Tamil Nadu, Assam, and West Bengal). The fitting parameters for all four states are summarized in Table 2.

**Figure 4:**
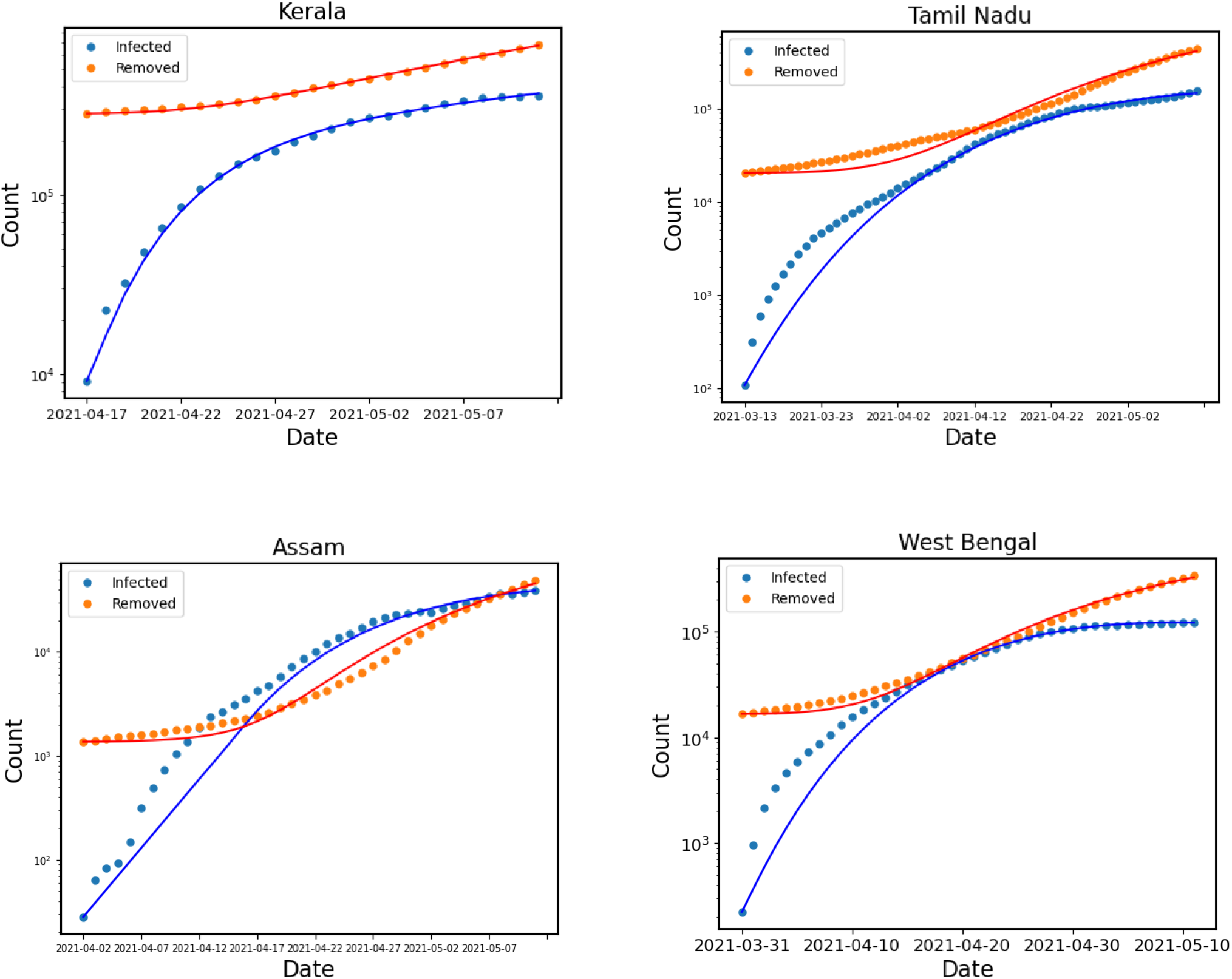
Blue and orange points indicated infected and removed individuals respectively for all election-bound states (Kerala, Tamil Nadu, Assam, and West Bengal). The fitting with the SIR case with time-dependent exponential *β* suppression model (as described in Section 2.4) was shown for all states with solid lines.

**Table 2:** A summary of the fitting parameters for the SIR case with time-dependent exponential *β* suppression model used to fit infected and removed individuals.

| Model | States | $\gamma, \beta_0, \alpha, \mu, t_l$ |
| --- | --- | --- |
| SIR with $\beta(t)$ exp | Kerala | 0.08, 0.70, 0.20, 0.20, 1.20 |
|  | Tamil Nadu | 0.12, 0.47, 0.22, 0.04, 0.91 |
|  | Assam | 0.09, 0.40, 0.19, 0.08, 13.97 |
|  | West Bengal | 0.13, 0.61, 0.14, 0.07, 1.23 |
|  | Whole election states | 0.11, 0.60, 0.20, 0.11, 0.11 |

In Figure 5, we showed the cumulative variation of infected and removed individuals for all four election-bound states. The Figure was similar to Figure 4, where the fitting with the SIR model with time-dependent exponential *β* suppression model was shown for cumulative variation of four election-bound states.

**Figure 5:**
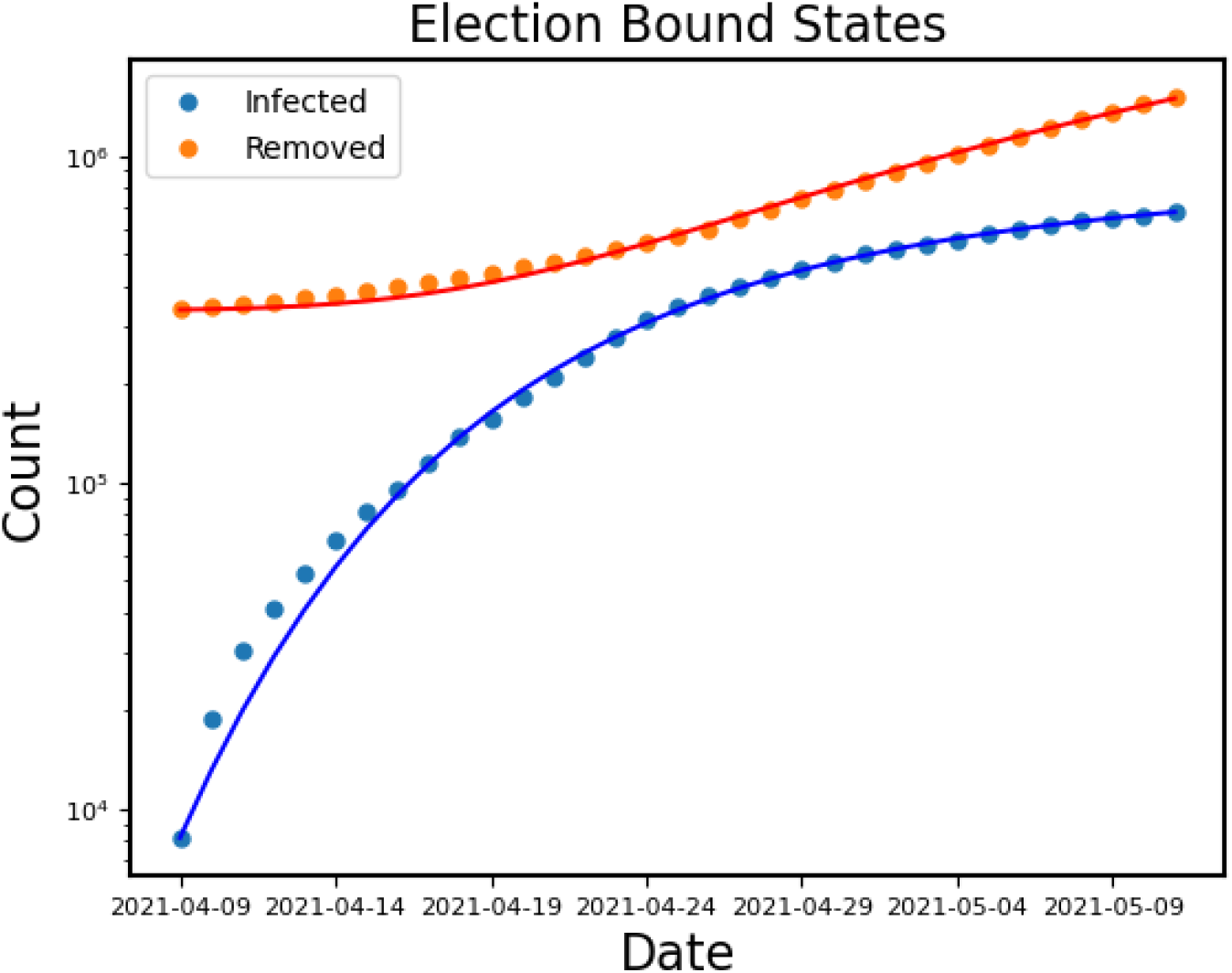
The same as in Figure 4 for the summation of all election-bound states in India.

We modeled the trend of the effective reproduction number for three epidemiological models (SIR, SIRD, and SEIR) using different mathematical functions. In Figure 6, ℝ(t) was fitted with different mathematical functions like Lorentzian, split Lorentzian, and Gaussian for SIR, SIRD, and SEIR models. We applied the least square method to fit ℝ(t). The top left of Figure 6 showed the evolution of R(t) during the election period for all the election-bound Indian states. ℝ(t) for the SIR model was well fitted with Lorentzian, split Lorentzian and Gaussian functions and the value of reduced chi-squared (*χ*^2^) was ∼0.05 for these functions. The right top of Figure 6 showed the variation of ℝ(t) for the SIRD model, which is fitted with the same three mathematical functions. ℝ(t) for the SIRD model was also well fitted with all the three functions and the value of reduced chi-squared (*χ*^2^) was ∼0.05 for these functions. The bottom of Figure 6 represented the variation of ℝ(t) for the SEIR model, which was well fitted with the Lorentzian function with reduced *χ*^2^ ∼4.79. The variation of ℝ(t) was also fitted well with a Gaussian function with a reduced *χ*^2^ value of ∼4.85.

**Figure 6:**
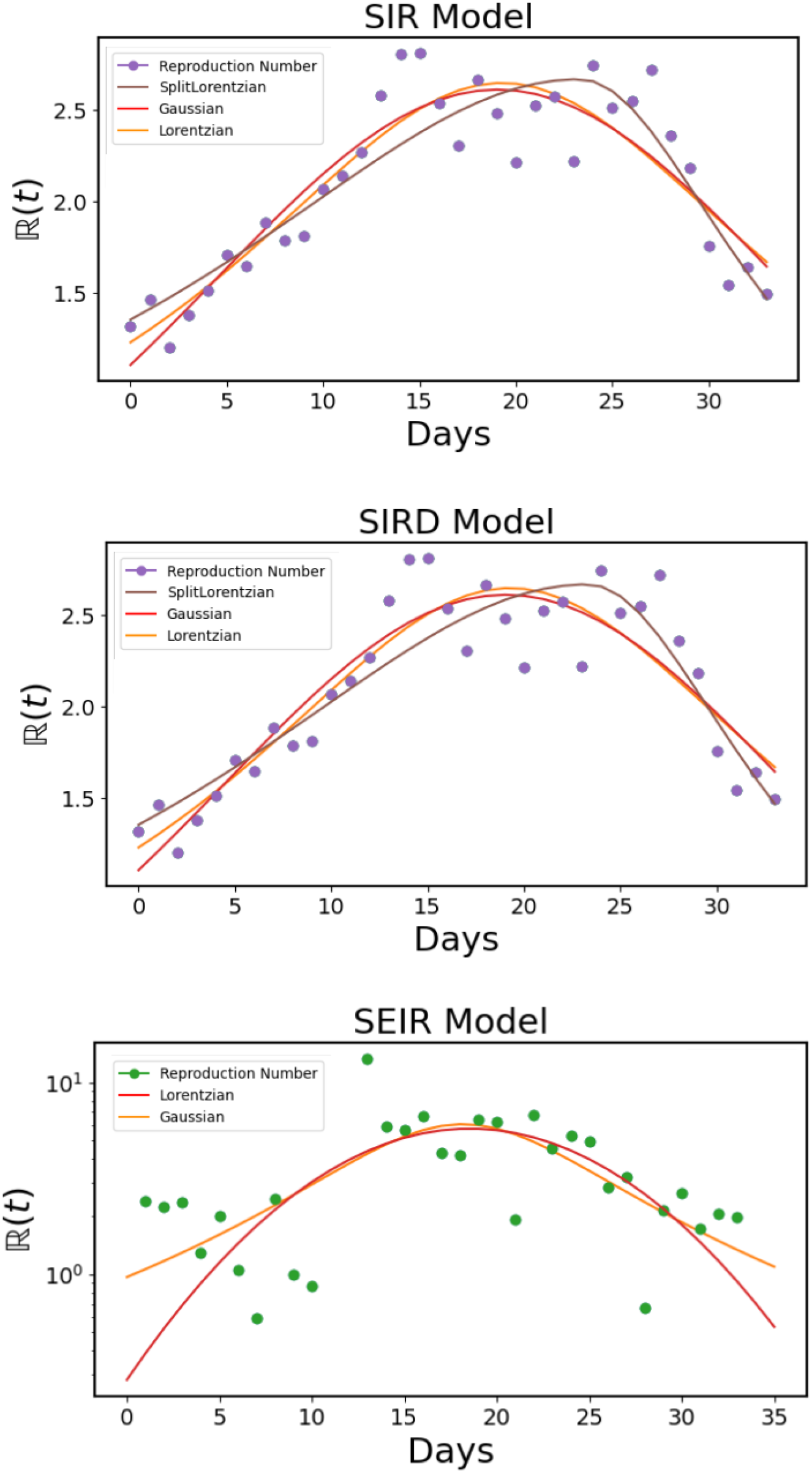
The variation of effective reproduction number (ℝ(*t*)) during election time is fitted with different mathematical functions using cumulative data for all election-bound states. The initial reference point represents the first date of 2021 Assembly Election (March 27, 2021).

## 5 Discussion

The impact of the assembly election on the transmission of Covid-19 was studied for all election-bound states separately. Most of the Indian states, where the election took place with multiple phases, were affected badly by Coronavirus.

Election campaigns, political activities including political rallies, meetings, overcrowded gatherings were held regularly for different cities of the states without maintaining proper Covid-19 protocol. These political events played a crucial role in a sharp rise in the number of infected individuals as well as in the significant increment in the effective reproduction number and contact rate of the Coronavirus.

The spread of Coronavirus depended on so many factors like social distance, personal hygiene, vaccination, environmental factors, etc. During the study, we assumed that these factors remained the same for these states, and the effect on the result is marginal. The drastic change in effective reproduction number of Covid-19, effective contact rate, and daily new cases during the election period implied that the bulk gathering during the political activities played a major role to spread the virus.

The result showed that for West Bengal and Assam, where the election took place in multiple phases, the new Coronavirus infected cases were increased unusually probably due to several political movements, meetings, gatherings during the prolonged pre-election, and election period. The comparative study of the variation of different fundamental parameters of the pandemic for all election-bound states and the rest of India suggested that the effect of election played a key role to increase the spread of the virus unusually. It was evident that the states where the election was completed in single-phase were comparatively less affected than the states where the election was conducted in multiple phases.

## 6 Conclusion

We have summarized the results of our study on the impact of political activities and elections on the Covid-19 spread rate for different Indian states during the March-May 2021 Assembly election using different mathematical models. Using epidemiological models such as SIR, SIRD, and SEIR, we investigated the effects of assembly election on the effective contact rate, and effective reproduction number. We looked at each election-bound state separately and found that all of them had a significant rise in effective contact rate and effective reproduction number during the election-bound period and immediately afterward, compared to the pre-election period. The impact of pre-election activities including political rallies, movements, and over-crowded gatherings was reflected clearly in the change of effective reproduction number. States with single-phase elections are comparatively less affected than the states where the election was conducted in multiple phases. From the first week of April 2021, the election commission imposed additional restrictions on large campaign rallies, meetings, and other political activities which helped to slow down the effective contact rate and the effective reproduction number in all election-bound states.

## Data Availability

We have used data from https://api.covid19india.org/12%E2%80%99 and https://github.com/datasets/covid-19

## Footnotes

1 https://www.scmp.com/news/china/society/article/3074991/coronavirus-chinas-first-confirmed-covid-19-case-traced-back

2 https://www.who.int/csr/don/05-january-2020-pneumonia-of-unkown-cause-china/en/

3 https://www.who.int/emergencies/diseases/novel-coronavirus-2019/technical-guidance/naming-the-coronavirus-disease-(covid-2019)-and-the-virus-that-causes-it

4 https://www.who.int/publications/m/item/weekly-epidemiological-update-on-covid-19-11-may-2021

5 https://www.worldometers.info/coronavirus/#countries

6 https://www.thehindu.com/news/national/india-to-start-covid-19-vaccination-drive-on-jan-16/article33536670.ece

7 https://timesofindia.indiatimes.com/india/covid-19-only-six-states-to-begin-vaccination-for-18-from-today/articleshow/82335939.cms

8 https://www.elections.in/upcoming-elections-in-india.html

9 https://eci.gov.in/files/file/12919-general-election-to-the-legislative-assemblies-of-assam-kerala-tamilnadu-west-bengal-and-puducherry-2021/

10 https://www.indiatoday.in/india/story/madras-hc-election-commission-covid-wave-officials-booked-murder-1795082-2021-04-26

11 https://api.covid19india.org/

12 https://github.com/datasets/covid-19

13 https://eci.gov.in/files/file/12919-general-election-to-the-legislative-assemblies-of-assam-kerala-tamilnadu-west-bengal-and-puducherry-2021/

14 https://www.census2011.co.in/census/state/puducherry.html#futurepop

15 https://www.census2011.co.in/census/state/kerala.html

16 https://www.census2011.co.in/census/state/tamil#nadu.html

17 https://www.census2011.co.in/census/state/assam.html

18 https://www.census2011.co.in/census/state/west+bengal.html

